# Patient Preferences for Diagnostic Imaging Services: Decentralize or not?

**DOI:** 10.1101/2024.03.18.24304499

**Authors:** Eline M. van den Broek-Altenburg, Jamie S. Benson, Adam J. Atherly, Kristen K. DeStigter

## Abstract

The objective of this study was to identify patient preferences for outpatient diagnostic imaging services and analyze how patients make trade-offs between attributes of these services using a discrete choice experiment (DCE).

We used a DCE with 14 choice questions asking which imaging locations patients would prefer. We used latent class analysis to analyze preference heterogeneity between different patient groups and to estimate the relative value they assign to different attributes of imaging services.

Our analysis showed that the “Experienced Patients” subgroup generally value diagnostic imaging services in both acute and chronic situations and had a strong preference for hospital outpatient radiology departments (HORD) that would provide services at lower costs, where their images would be interpreted by a specialty radiologist, the clinic would be recommended by their PCP, online scheduling would be available, service rating were higher, and travel and wait times would be shorter. New Patients significantly valued the service rating of the hospital outpatient radiology departments (HORD) and online scheduling.

HORDs can be more competitive by providing services that live up to expectations better than available retail radiology clinics (RRCs). Most RRCs do not currently offer online scheduling so ease of use may also steer patients towards HORDs. HORDs have the advantage of being linked to the main medical center which has the reputation of having clinical expertise and more sophisticated technology. We conclude that there is room for medical centers to build HORDs that provide an appealing and competitive alternative to current RRC.

## Introduction

An important challenge for many hospitals and clinics is addressing the escalating diagnostic imaging demands for hospital outpatient radiology departments (HORDs)^1^. HORDs have struggled to keep up with the increasing need for imaging, with MRI and CT imaging rates for adults more than doubling from 2006 to 2016 ^2^. This escalating demand for services, coupled with an increase in patient complexity not only leads to increased costs, but also reduced accessibility and quality. Delays in imaging at HORDs can also lead to delayed care delivery, which can negatively impact patient outcomes.

Retail radiology clinics (RRCs) emerged from the general retail health clinics’ market boom as a potential solution to the imaging bottleneck effect of HORDs. RRC offer a variety of diagnostic imaging services, typically at lower cost, at a decentralized clinic ^3-5^. RRCs have gained popularity by cutting wait-times, increasing accessibility, and reducing costs ^3-5^. Despite the increase in RRC utilization, price variability and unreliable quality of the technology used, imaging interpretation and reporting of images have led RRCs to be labeled as low-value substitutes to HORD services ^6-8^. For example, one study found significant variability in diagnostic reports from 10 different RRCs performing an MRI scan of the lower back on the same patient within a 3-week period 5. Another study reported that radiology departments in tertiary care centers are frequently asked to perform secondary interpretations of imaging studies, finding that discrepancy rates vary widely ^6^.

In parallel, hospital networks are struggling with care integration and coordination, as well as maintaining quality standards across all departments and campuses within their network. Some hospital systems are now looking at options to develop their own RRCs to provide convenient and efficient locations, without sacrificing the technological or interpretation quality, while leveraging the existing trust patients may have with their hospital system. This is a potentially valuable innovation for patients, as they would be able to receive the “specialty read” quality at locations with shorter wait times and lower out-of-pocket costs.

To successfully decentralize imaging services at a larger scale, however, it is important to define value not just from the perspective of the healthcare system, but also from the perspective of the patient. Despite widespread encouragement to incorporate the patient voice in healthcare, developments such as RRCs are usually done without meaningful input from patients ^7^. In the push for patient-centeredness, CMS recommends hospital departments incorporate patient insight into their service quality measures ^8^.

Existing research on patient preferences in diagnostic imaging services is sparse, however, and what exists is often contradictory ^9-15^. Though survey studies have explored from whom patients prefer to learn their results and how (e.g., by phone, e-mail, or in-person), findings to date have been inconsistent ^9–11,16^. For example, one study found a majority preference for learning results in-person and from a radiologist^10^ while another found that most patients prefer to learn results from their primary care provider, over the phone ^11^.

The objective of this study is to identify patient preferences for outpatient diagnostic imaging services and analyze how patients make trade-offs between attributes of these services using a stated choice experiment. In this study, we analyze patient’s individual preference heterogeneity for these services and study how preferences vary among and within different patient populations.

## Methods

### Discrete Choice Experiment

In this study, we used a Discrete Choice Experiment (DCE) to explore which attributes of imaging services matter to patients and how they make trade-offs between these attributes. A DCE provides the opportunity to estimate pair-wise choices and analyze marginal values for the total value of a health service or good ^17^. The unique contribution of a DCE is that it allows researchers to analyze the trade-offs that patients are willing to make, including options that may not currently exist but could in the future. We do this by using a survey in which patients answer a repeated set of choice tasks. In these choice tasks, they are asked to choose their preferred option of three locations offering imaging services. The imaging locations vary by different attributes and levels of those attributes. We explain this further in the paragraph about the experimental design.

With the DCE, we can estimate the utilities that patients assign to different attributes of imaging services. Utilities represent the total satisfaction or benefit derived from consuming a good or service and the capacity of a service to satisfy patients’ wants. To estimate the relative utilities patients assign to different aspects of services, the DCE included 14 choice tasks. The experimental design of the DCE was based on “prior” estimates of the utilities for attributes of the choice which are calculated by using expectations on what the model parameters will be. These numbers are based on sample data from a pilot and from the literature.

### Qualitative Research

To understand which aspects or attributes of imaging services are important to patients and should therefore be included in the choice experiment, we first performed qualitative research to identify key aspects of value to patients. This qualitative work defined the most important attributes of imaging services to patients and provided the base for a DCE. We performed two focus groups, each with 12 patients over the age of 18 who had received outpatient imaging services in 2019 from a medium-sized academic medical center, at the main campus of a six-hospital, two-state network in the Northeastern part of the United States. A question guide aimed at understanding how patients perceive their radiology experiences led participants through their focus group session. Questions were intended to be exploratory and were sometimes followed by probes to allow differences between patient insights and experiences to emerge. From the thematic analysis of the focus groups, we learned that important themes included: overarching trust in system and referring providers; preference for transparent and informative communication; personal interaction and compassionate bedside manner from staff; and accessibility of radiology services and the facility. We also performed qualitative research among leaders in the radiology department of the medical center who were asked what they considered to be important to patients. Their suggestions provided additional input for the stated choice design, such as the potential importance of wait time to be seen and whether or not the location for getting imaging was recommended by patient’s primary care provider.

### Experimental Design

Two discrete choice experiments were designed to represent an acute patient’s need (X-ray service), and a sub-acute need using Magnetic Resonance Imaging (MRI service). Participants were asked to imagine a situation where they were hurt and needed imaging services. Participants were randomized between two arms with the following descriptions of the choice situation:

- *Situation 1): “For the purpose of this study, suppose you hurt your arm and your primary care provider wants to send you for an X-ray. You have three options of locations where you can have your imaging done*.*”*
- *Situation 2): “For the purpose of this study, suppose you hurt your back a while ago, and are having persistent pain. Your primary care provider wants to send you for an MRI. You have three options of locations where you can have your imaging done*.*”*

The DCE included 14 choice tasks where patients were asked to choose the imaging clinic they preferred. Each choice task had three different clinics which varied by 10 different attributes, which were determined by prior qualitative research. These selected attributes included: whether the interpreting radiologist is a general or sub-specialty radiologist; whether the clinic was recommended by their primary care physician (PCP); time to results; out of pocket cost; wait time to an appointment; travel time to the clinic; parking costs; parking accessibility; service; and whether or not online scheduling is available. Service is a multifactorial attribute (e.g., staff attentiveness and facility amenities) combined into a star rating. The rating scale is between one and five stars, with a five-star rating representing an excellent service and a one-star rating suggesting a poor service, as rated by other hypothetical patients. Figure 1 shows an example of a choice task.

**Figure 1.**
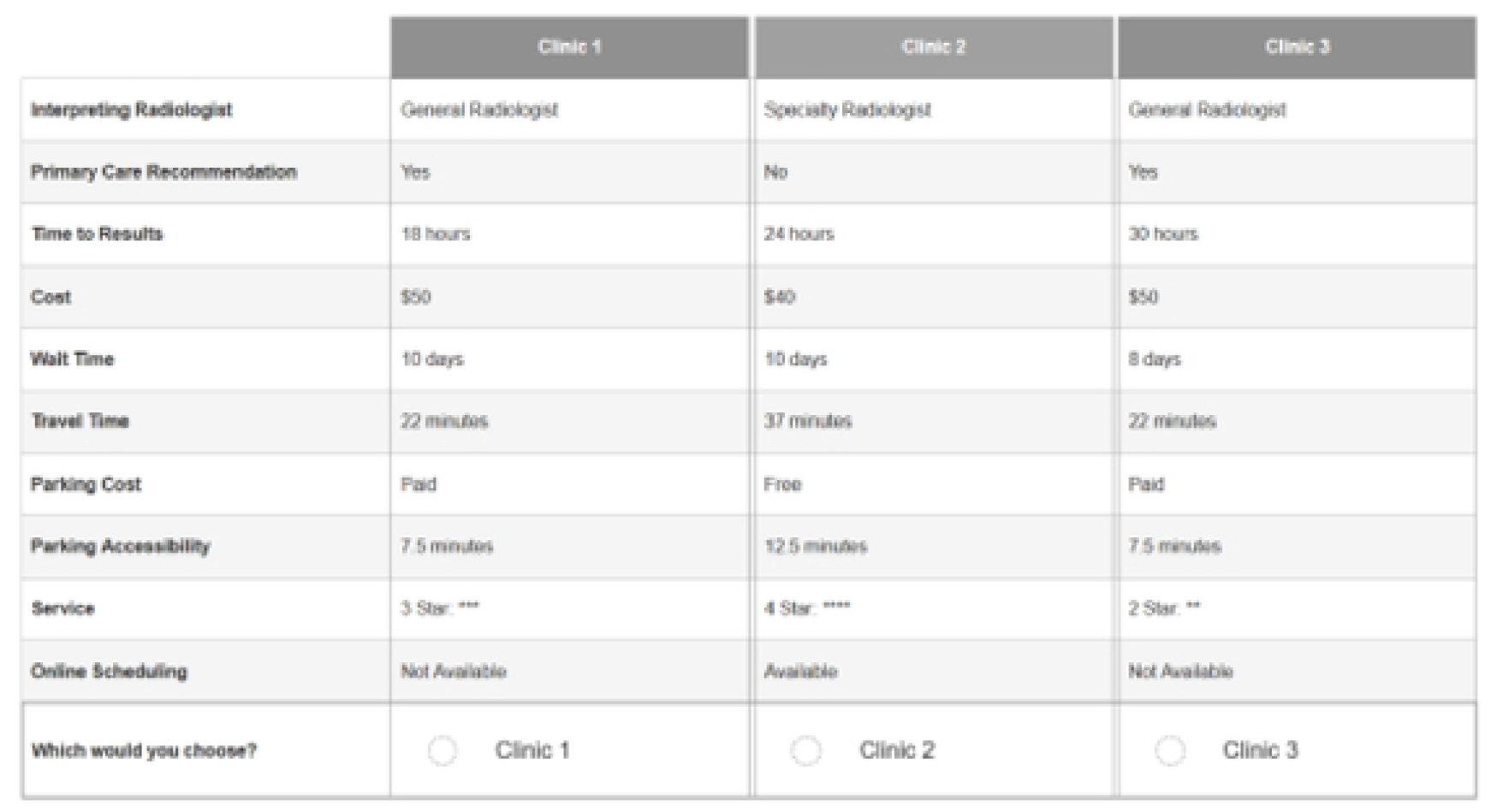
Example ofa Choice Task in the DCE

Following the DCE choice tasks, we asked patients attitudinal questions on a 1–5-point Likert scale from “strongly disagree” to “strongly agree”. These questions were borrowed from and validated by the national Medical Expenditures Panel Survey (MEPS) ^19^, a set of large-scale surveys of families and individuals, their medical providers, and employers across the United States. The questions focused on perceived need for healthcare, need for insurance, risk-aversion, and perceived personal health status.

### Data Source, Participants and Study Size Estimation

Our data were sampled from an online Centiment panel between April 11, 2021 through November 19, 2021, with a second sample between May 16, 2022, and June 24, 2022. Centiment contacted approximately 472 individuals in the catchment area of a moderate size academic medical center in a rural Northeastern part of the United States. Participants completed written consent before continuing to the online survey questions. All data were fully anonymized before shared with the study team. The Institutional Review Board at the University of Vermont reviewed the study and determined it was exempt from full review.

Of the total sample, 268 finished the survey and met initial inclusion criteria (Age >= 18): 134 were assigned to the arm X-ray group, 134 to the back MRI group. We excluded 98 subjects for failing consistency criteria, qualifying for a closed quota, failing a bot-behavior check, or failing an attention (response quality) check, leaving a final sample of 170: 84 in X-ray, 86 in MRI. As each subject answered 14 choice tasks, we obtained an effective sample size of n=2,380 for modeling. We used NGene 1.2.1 (ChoiceMetrics, 2018) to estimate the minimum sample size required for this study.

### Statistical Methods

We used a mixed logit model (ML) to estimate the probability of a choice alternative being chosen, depending on the characteristics of the choice (attributes and levels) and the characteristics of the chooser. We used latent class analysis (LCA) which addresses the issue of unobserved preferences of patients by probabilistically segmenting a sample population into different groups or “classes” based on a latent variable^24^. Class membership is first defined by a membership function including the indicator variables, after which the utility functions of different classes can be estimated. All statistical analyses were conducted using Stata 18 (StataCorp LLC, 2023).

### Latent Class Analysis

The latent class conditional logit (LCL) model further extends the ML model by incorporating a discrete representation of unobserved preference heterogeneity within groups in a population. The LCL model might explain, for example, that patients who had previous imaging services are more likely to fall into the class that is more sensitive to appointment wait time, while older patients might be more likely to fall into the class that is more sensitive to PCP recommendation.

## Results

### Descriptive Results

A total of 84 patients answered questions about preferences for attributes of an X-ray; 86 people responded to the MRI choice questions. The summary statistics are reported in Table On average, patients answering choice questions on X-ray tended to be female (60%), white (91%) and live in rural areas (58%); about half had private insurance (46%) and relatively few had met their insurance deductible (21%). Patients answering the choice questions on the MRI were generally similar, with 67% female, 96% white, 69% in rural areas, 45% with private insurance, 22% had met their insurance deductible. Patients receiving the X-ray choice questions had had an average of 4.8 previous images while MRI patients had had 2.9.

### Mixed logit Results

The results of the mixed logit model are shown in Table 2 where we separated results for X-ray and MRI. We found that out-of-pocket costs, interpreting doctor specialty, whether or not the clinic was recommended by the primary care provider, the wait time to results, the clinic service rating and online scheduling were all statistically significant and had the expected signs for both MRI and X-ray. Patients were less likely to choose a clinic if the out-of-pocket costs were higher and the wait time to results was longer, but more likely to choose it if images were interpreted by a specialty radiologist, the clinic was recommended to them by their primary care provider, the service rating was higher and if online scheduling was available. For X-ray, free parking was associated with a higher probability of choosing a clinic.

**Table 1:** Descriptive Statistics

|  | **X-ray** |  | **MRI** |  |
| --- | --- | --- | --- | --- |
| Age (mean, sd) | 46.0 | (16.1) | 47.8 | (16.9) |
| Income (USD) (mean, sd) | \$48,425 | (\$40,939) | \$47,444 | (\$47,881) |
| Number of Previous Images (mean, sd) | 4.8 | (1.5) | 2.9 | (1.7) |
| Agreement: I don't need insurance (mean, sd) | 1.5 | (0.9) | 1.7 | (1.2) |
| Agreement: Insurance isn't worth the cost (mean, sd) | 2.3 | (1.3) | 2.4 | (1.4) |
| Agreement: I take more risks than others (mean, sd) | 2.3 | (1.2) | 2.3 | (1.2) |
| Agreement: I can fight illness without a doctor (mean, sd) | 2.5 | (1.3) | 2.4 | (1.2) |
| Agreement: I am healthier than others (mean, sd) | 2.7 | (1.1) | 2.7 | (1.1) |
| Gender Identity (% , n) |  |  |  |  |
| Male | 38.1% | 32 | 32.6% | 28 |
| Female | 59.5% | 50 | 67.4% | 58 |
| Non-Binary | 2.4% | 2 | 0.0% | 0 |
| Race (% , n) |  |  |  |  |
| White | 91.7% | 77 | 96.5% | 83 |
| Black | 1.2% | 1 | 0.0% | 0 |
| Asian | 2.4% | 2 | 2.3% | 2 |
| AIAN | 3.6% | 3 | 1.2% | 1 |
| Multiple | 1.2% | 1 | 0.0% | 0 |
| Ethnicity (% , n) |  |  |  |  |
| Not Hispanic | 92.9% | 78 | 96.5% | 83 |
| Hispanic | 7.1% | 6 | 3.5% | 3 |
| Rurality (% , n) |  |  |  |  |
| Urban | 14.3% | 12 | 14.0% | 12 |
| Suburban | 27.4% | 23 | 17.4% | 15 |
| Rural | 58.3% | 49 | 68.6% | 59 |
| Education Level (% , n) |  |  |  |  |
| Less than High School | 2.4% | 2 | 1.2% | 1 |
| High school | 27.4% | 23 | 23.3% | 20 |
| Some college, no degree | 25.0% | 21 | 17.4% | 15 |
| Associate's: vocational | 0.0% | 0 | 3.5% | 3 |
| Associate's: academic | 6.0% | 5 | 2.3% | 2 |
| Bachelor's (BA, AB, BS, BBA) | 23.8% | 20 | 32.6% | 28 |
| Master's | 11.9% | 10 | 15.1% | 13 |
| Professional, Doctoral | 3.6% | 3 | 4.7% | 4 |
| Insurance Status (% , n) |  |  |  |  |
| Insured - Private | 46.4% | 39 | 45.3% | 39 |
| Insured - Medicare | 21.4% | 18 | 24.4% | 21 |
| Insured - Medicaid | 26.2% | 22 | 23.3% | 20 |
| Uninsured | 6.0% | 5 | 7.0% | 6 |
| Deductible Status (% , n) |  |  |  |  |
| Met | 21.4% | 18 | 22.1% | 19 |
| Unmet | 44.0% | 37 | 45.3% | 39 |
| No Deductible | 34.5% | 29 | 32.6% | 28 |
| Health Status (% , n) |  |  |  |  |
| No Chronic Condition | 50.0% | 42 | 47.7% | 41 |
| Chronic Condition | 50.0% | 42 | 52.3% | 45 |

**Table 2:** Results of the MMNL Model

|  | (MRI) | (MRI) | (MRI) | (X-ray) | (X-ray) | (X-ray) |
| --- | --- | --- | --- | --- | --- | --- |
|  | B | SE | 95% CI | B | SE | 95% CI |
| Out of Pocket Cost | -0.004*** | (0.001) | -0.005,-0.003 | -0.006* | (0.002) | -0.010,-0.001 |
| Interpreting Doctor Specialty | 0.732*** | (0.116) | 0.504,0.960 | 0.374** | (0.122) | 0.135,0.614 |
| Clinic Recommended by PCP | 0.626*** | (0.112) | 0.407,0.845 | 0.678*** | (0.155) | 0.374,0.982 |
| Wait Time to Results | -0.012* | (0.006) | -0.024,-0.001 | -0.013*** | (0.004) | -0.020,-0.006 |
| Travel Time to Clinic | -0.007 | (0.006) | -0.004,-0.018 | -0.018* | (0.008) | 0.001,0.034 |
| Wait Time to Appointment | -0.014 | (0.018) | -0.050,-0.022 | -0.023* | (0.012) | 0.000,0.047 |
| Clinic Service Rating | 0.520*** | (0.096) | 0.332,0.708 | 0.451*** | (0.126) | 0.205,0.698 |
| Free Parking | 0.150 | (0.097) | -0.040,0.339 | 0.669*** | (0.108) | 0.457,0.881 |
| Clinic Parking Lot Distance | -0.041 | (0.023) | -0.087,0.005 | -0.011 | (0.007) | -0.025,0.002 |
| Online Scheduling Availability | 0.288* | (0.112) | 0.068,0.508 | 0.375*** | (0.089) | 0.201,0.550 |
| *** p<.001, ** p<.01, * p<.05 |  |  |  |  |  |  |

The attributes that mattered the most to patients for both MRI and X-ray were specialty radiologist reading (0.732, standard error= 0.116 for MRI; 0.374, se =0.122) for X-ray); recommendation by the primary care provider reading (0.626, se=0.112 for MRI; 0.678, se=0.155 for X-ray) and the clinic’s service rating reading (0.520, se=0.096 for MRI; 0.451, se=0.126 for X-ray).

### Latent Class Analysis

Table 3 shows the results of the Latent Class model where age, gender, education, income, whether someone had more than two previous scans, whether they considered themselves healthier than others, whether they had private insurance and whether they were more likely to take risks than others were the indicator variables estimating the probability of class membership. We found that 54.7 percent of patients were in class 1 and 45.3 percent in class 2.We found that females, older patients, whether they had at least two previous scans and those who were less likely to take risks than others were highly predictive of being in class 2 (p<0.02) which we therefore labeled as the “Experienced Patients” class. Those who had fewer than two previous scans were more likely to be in class 2 (2.7111, p<0.01) as were male patients (−0.8479, p=0.02) and younger patients (−1.7197, p=0.02), which we labeled “New Patients”. The results show that someone who had a 1-point increase in the Likert scale for “more likely to take risks than others” was significantly more likely to be in class 2 (−3.1354.7277, p=0.01). they offer at the centralized location.

**Table 3:** Results of the Latent Class Analysis

|  | **Class membership** | **Standard error** |
| --- | --- | --- |
| Male | -0.84793** | (.3791) |
| Income (ref: lowest income category) | 0.1477 | (.3979) |
| Age (years) | 1.7196** | (.7559) |
| More than 2 previous scans | 2.7110*** | (.9873) |
| Education (ref: no high school) | -12.9818 | (3.6767) |
| More likely to take risk than others | -3.1353*** | (1.2389) |
| Private insurance | -1.0671 | (1.1982) |
| Healthier than others | -14.4012 | (10.5153) |
|  | **Class 1 (Experienced Patients) B (SE)** | **Class 2 (New Users) B (SE)** |
| Costs > current | -0.1774*** (0.0367) | -0.2783 (0.1764) |
| Specialty read | 0.35123*** (0.0419) | 0.0525 (0.2753) |
| PCP recommendation | 0.3890*** (0.0432) | 0.1641 (0.2178) |
| Online Scheduling | 0.1240*** (0.0419) | 4.7541*** (0.4703) |
| Service rating | 0.3033*** (0.0509) | 0.1148*** (0.2636) |
| Time to Results | -0.0225 (0.0340) | -0.0289 (0.1763) |
| Travel Time | -0.1473*** (0.0324) | -0.2649 (0.1618) |
| Wait Time | -0.1165*** (0.0447) | -0.1271 (0.1968) |
| Access to parking | -0.0308 (0.0254) | -0.0509 (0.1829) |

\*\*\* $p < .01$ , \*\* $p < .05$ , \* $p < .10$

When decentralizing some services away from the main hospital, the focus should be on passing on the medical center’s high service rating to the HORD and offering online scheduling. Indeed, patients value easier access, shorter wait times and lower out-of-pocket costs which the growing popularity of RRCs has shown. HORDs can win in popularity by making sure they attain and retain high star ratings while offering better access than RRCs. Primary care providers can potentially play an important role in directing patients to HORDs for their diagnostic imaging services.

For patients in class 1 (“Experienced Patients”), costs (−0.1774), specialty read (0.3512), PCP recommendation (0.3890), travel time (−0.1474), wait time (−0.1165), service (0.3033) and online scheduling (0.1240) all had significant effects (p<0.01) and had the expected signs. For example, patients in the “Experienced Patients” class will be more likely to choose a clinic location if it is lower cost, takes less travel time, has shorter wait times for results, if results are read by a specialty radiologists, the clinic is recommended by their PCP, service is better and online scheduling is available. Patients in the “New Patients” class only cared about online scheduling (4.7542) and service rating (1.1149), but the effect size was relatively high. None of the other attributes of the service would affect their choice.

### Marginal Rates of Substitution

Table 4 shows an analysis of the trade-offs patients were willing to make, known as the marginal rates of substitution. We found that for patients in the Experienced Patient class, even though most attributes significantly affected their choice for clinic, the effect size was considerably smaller than for the New Patients class. Experienced Patients were willing to pay: $2 more than what they currently pay (out-of-pocket) to have their images read by a radiologist specialist; $2 more to go to a clinic that was recommended by their PCP; $0.70 more for online scheduling; $1.70 more for a 1-point higher star rating; $0.80 more to have a clinic that would be 1 minutes closer than their current one; $0.65 more for a clinic that had a 1-hour shorter wait and $0.17 more for a clinic that would decrease their walk-up time by 1 minute. The New Patients were willing to pay $17 more for online scheduling and $4 more for a higher star rating.

**Table 4:** Willingness-to-Pay (MRS)

| **WTP for Attribute (\$\$)** | **Class 1**<br>**(Experienced Patients)** | **Class 2**<br>**(New Patients)** |
| --- | --- | --- |
| Radiologist Specialty | 1.980 | 0.189 |
| PCP Recommendation | 2.192 | 0.590 |
| Time to Results | -0.127 | -0.104 |
| Travel Time to Clinic | -0.831 | -0.952 |
| Wait Time to Appointment | -0.657 | -0.457 |
| Service Quality | 1.709 | 4.005 |
| Parking Accessibility | -0.174 | -0.183 |
| Online Scheduling Available | 0.699 | 17.079 |

### Attribute Importance

Patients were also asked to rank-order the attributes in how they prioritized them when making a choice for a clinic. This is usually consistent with the magnitude of the coefficients in the choice models, but also allows us to see if there was any attribute non-attendance. This means that when processing the attributes, some patients may not consider particular attributes at all. The violin plot in Figure 2 shows the results of the question on attribute importance. We see that the results are consistent with the ML results: the most important attributes included interpreting doctor specialty level, PCP recommendation, and costs. There was a small difference between the X-ray and MRI arms: among X-ray patients, service rating and parking access also ranked highly.

**Figure 2.**
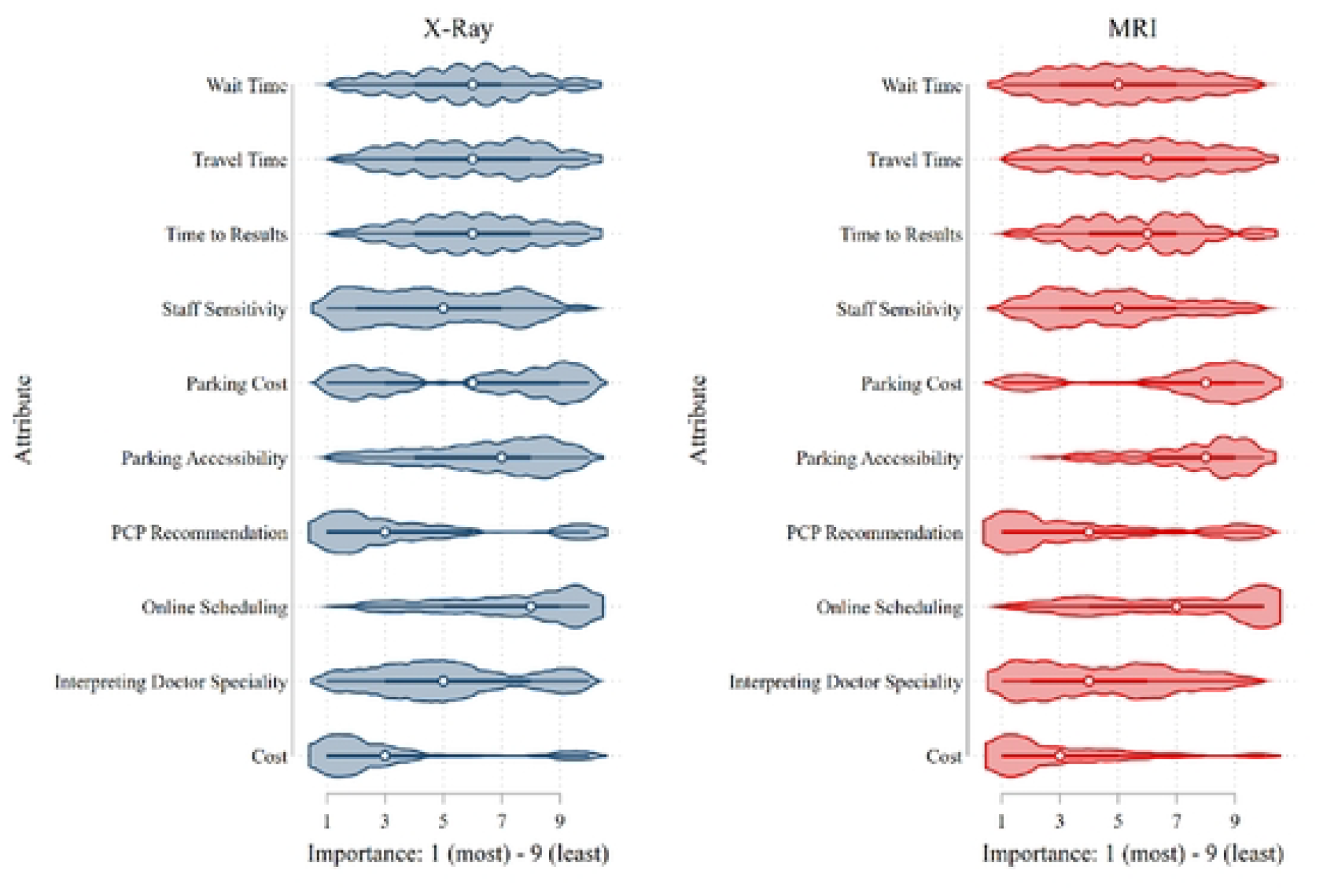
Attribute Importance

## Discussion

In this paper, we sought to identify patient preferences for outpatient diagnostic imaging services. We analyzed how patients make trade-offs between attributes of services using a discrete choice experiment. We explored patient’s individual preference heterogeneity for these services and reported how preferences vary among and within different groups of patients.

In our base analysis, we found that specialty reading of images, PCP recommendation, lower costs, travel time and wait time, as well as higher star rating (representing better service or reputation) and online scheduling are all significant predictors of choice regarding where to get diagnostic imaging services. However, when we segmented the sample population deterministically, we found that males, younger people and more likely to take risks than others only We termed this group “New Patients” as they had significantly fewer previous scans and generally do not highly value health services. Insurance status, health status and chronic conditions, education and income did not define class membership.

These results suggest that decision-makers looking to decentralize imaging services while incorporating patient preferences for attributes of those services should differentiate between the different patient sub-populations they are serving. This requires careful consideration of patient characteristics as well as preferences. Overall, we found in this study that New Patients care about the reputation or star rating of a clinic and online scheduling availability. But additionally, Experienced Users – who will be the majority of radiology users – are focused on wait time, price and recommendations from primary care providers. This is a concrete message to medical centers seeking to decentralize their services away from the main hospital: patients will want to know that the new location offers the same service level in addition to convenience and will be relying on their primary care providers for advice, suggesting outreach to primary care providers will be important for success.

### Limitations

Although Centiment used a quota sampling approach, the gender balance may not be reflective of the total population, although we do not believe this is a major threat to external validity. However, while our sample is largely representative of the larger population, we measure intention for hypothetical choices and cannot say for sure that these consistently translate to real-life behavioral trade-offs, especially in acute situations. More work needs to be done to further explore factors that affect decision-making and preferences in these circumstances.

## Conclusions

In this study, we analyzed the trade-offs patients make between attributes of radiology services to inform decision-making around designing optimal HORDs. Our analysis showed that a patient population can and should be segmented into subgroups that evaluate the value of imaging services differently. The “Experienced Patients” subgroup generally value diagnostic imaging services in both acute and more chronic situations and they had a strong preference for a HORD that would provide services at lower costs, where their images would be interpreted by a specialty radiologist, the clinic would be recommended by their PCP, online scheduling would be available, service rating were higher, and travel and wait times would be shorter. HORDs can therefore be more competitive by providing services that live up to these expectations better than available RRCs.

New Patients significantly valued the service rating of the HORD and online scheduling. Most RRCs do not currently offer online scheduling so ease of use may also steer these potential future patients towards HORDs. More importantly, HORDs have the advantage of being linked to the main medical center which has the reputation of having clinical expertise and more sophisticated technology. We conclude that there is room for medical centers to build HORDs that provide an appealing and competitive alternative to the current RRC. RRCs have the risk of unreliable quality in the technology and imaging interpretation and it is therefore desirable that HORDs provide the same or more benefits while maintaining the quality care

## Data Availability

GitHub

https://github.com/ElinevandenBroek/

